# Monitoring One Heart to Help Two: Heart Rate Variability and Resting Heart Rate using Wearable Technology in Active Women Across the Perinatal Period

**DOI:** 10.1101/2022.04.22.22274195

**Authors:** Shon P. Rowan, Christa L. Lilly, Elizabeth A. Claydon, Jenna Wallace, Karen Merryman

## Abstract

**BACKGROUND:** Characterizing normal heart rate variability (HRV) and resting heart rate (RHR) in healthy women over the course of a pregnancy allows for further investigation into disease states, as pregnancy is the ideal time period for these explorations due to known decreases in cardiovascular health. To our knowledge, this is the first study to continuously monitor HRV and RHR using wearable technology in healthy pregnant women.

**METHODS:** A total of eighteen healthy women participated in a prospective cohort study of HRV and RHR while wearing a WHOOP® strap prior to conception, throughout pregnancy, and into postpartum. The study lasted from March 2019 to July 2021; data were analyzed using linear mixed models with splines for non-linear trends.

**RESULTS:** Eighteen women were followed for an average of 405.8 days (SD=153). Minutes of logged daily activity decreased from 28 minutes pre-pregnancy to 14 minutes by third trimester. A steady decrease in daily HRV and increase in daily RHR were generally seen during pregnancy (HRV Est. = -0.10, *P*<0.0001; RHR Est. = 0.05, *P*<0.0001). The effect was moderated by activity minutes for both HRV and RHR. However, at 49 days prior to birth there was a reversal of these indices with a steady increase in daily HRV (Est.=0.38, *P*< 0.0001) and decrease in daily RHR (Est. = -0.23, *P*< 0.0001), regardless of activity level, that continued into the postpartum period.

**CONCLUSIONS:** In healthy women, there were significant changes to HRV and RHR throughout pregnancy, including a rapid improvement in cardiovascular health prior to birth that was not otherwise known. Physical activity minutes of any type moderated the known negative consequences of pregnancy on cardiovascular health. By establishing normal changes using daily data, future research can now evaluate disease states as well as physical activity interventions during pregnancy and their impact on cardiovascular fitness.

## Introduction

Despite data that indicate a myriad of benefits of activity during pregnancy^1^, qualitative evidences suggests that there are consistent cultural expectations that pregnant women “should sit down and slow down”.^2^ These cultural expectations seem to be internalized with only 3-15% of those pregnant meeting current physical activity guidelines compared to 24-26% of non-pregnant individuals.^3^ Currently, based on clear evidence that physical activity and exercise in pregnancy is safe, the American College of Obstetrics and Gynecology (ACOG) recommends that women with uncomplicated pregnancies initiate or continue physical activity.^1^ They also advise that women who intend to get pregnant but do not have healthy lifestyles should focus on adopting them when possible prior to pregnancy. However, pregnant woman seeking advice from an obstetrician regarding physical activity in pregnancy may still receive conflicting answers. In a survey by Bauer et al., the majority of physicians surveyed were not familiar with the most recent ACOG guidelines.^4^ While the establishment of guidelines regarding the safety of physical activity in pregnancy has been agreed upon by leading organizations,^5^ many studies of exercise in pregnancy rely on retrospective data collected postpartum.

Wearable technology is an increasingly popular area of fitness tracking and in vivo data collection that opens doors for novel data collection in the area of pregnancy fitness.^6^ The WHOOP® strap (Strap 2.0; WHOOP, Inc., Boston, MA, USA) is a commercially available wearable device that provides continuous physiologic data monitoring and training recommendations based on proprietary scientific research.^7^ WHOOP® uses heart rate variability (HRV), along with resting heart rate (RHR) and sleep patterns to determine readiness for activity.^8^ HRV measures the irregularity of heart beat rhythm over time and is considered a low-cost, noninvasive measurement of overall competence of the autonomic nervous system.^9^ RHR measures the number of times a heart beats every 60 seconds at rest and is widely regarded as a measure of cardiovascular fitness.^10^ Combined, higher HRV and lower RHR represent higher levels of fitness. When evaluated together, HRV and RHR provide important insights regarding fitness and recovery levels of athletes.^11^

Previous studies have relied on intermittent measurement of autonomic responses using 24-hour holter monitoring or shorter HRV recordings.^12,13^ Using 18-minute HRV recordings at 28, 32 and 36 weeks, May et al. found that regardless of maternal exercise, heart rate increased throughout pregnancy.^12^ However, exercise during pregnancy resulted in lower maternal RHR and increased HRV, which indicate improvements in autonomic control.^12^

Limited research has started to use continuous monitoring of different physiological variables with wearable technology during pregnancy.^6, 14, 15^ However, these are limited by short length of monitoring, lack of monitoring of both HRV and RHR, and/or limited participants (case study).^6^ This is a burgeoning field of study and the potential to increase the time of follow-up during the perinatal period, measure HRV and RHR, and include an adequate sample of participants provides the potential to inform future research and current clinical practice.

The current study is designed to explore and describe the autonomic response in physically active women prior to, during, and after pregnancy when monitored in a continuous fashion. Continuous monitoring of physiologic data provides a novel view into cardiovascular workload and capacity during pregnancy. We had two main goals for this study. Our first aim was to use descriptive analyses to examine HRV and RHR changes during pregnancy in a sample of healthy women who were physical activity at least three days a week prior to pregnancy. Our second aim was to understand the relationship between increased activity during pregnancy and variations in HRV and RHR. We hypothesized that increasing activity would positively impact changes in HRV and RHR during pregnancy.

## Methods

### Recruitment

Following Institutional Review Board approval at West Virginia University, a total of 38 women were enrolled from March 2019 until August 2020. Inclusion criteria included women ages 18-35 years old, who were currently physically active at least three or more times per week. For inclusion, women also had to not be currently pregnant, but hoping to conceive within the next six months. Recruitment occurred primarily through the Reproductive Endocrinology and Infertility clinic at West Virginia University (WVU). Data were collected between March 2019 and July 2021.

### Wearable Device

Participants were given a WHOOP® Strap 2.0 (WHOOP, Inc., Boston, MA, USA) and asked to wear it continuously on their non-dominant arm from enrollment and throughout pregnancy, delivery, and postpartum. The WHOOP® devices were purchased with internal research funds through the WVU Department of Obstetrics and Gynecology. This strap transmitted continuous data to the participants smartphone and to a WHOOP cloud platform. Participants were able to see their daily physical activity information on their phones. The comprehensive data from all participants was then downloaded from the WHOOP cloud platform for analyzsis.

### Monthly Survey

A monthly survey (See Appendix) was sent via email asking participants about their continued participation as well as any changes in exercise (e.g. “How many days per week do you typically exercise?”), medical history (e.g. “Do you have any medical conditions” and “Will these medical conditions affect your ability to exercise while pregnant?”), pregnancy history (e.g. “How many times have you been pregnant?”), and pregnancy status (e.g. “When do you plan to start attempting conception?”). This survey allowed for participant retention, feedback, and to gauge overall health and fitness. The monthly response rate ranged from 89-100%, indicating excellent retention.

### Measures

Data were imported from WHOOP® in three different tables: 1) daily HRV and RHR; 2) recorded activity per participant, including time in each heart rate zone; and 3) a daily output of strain.

*Cardiovascular fitness* was assessed with two daily measures: HRV and RHR. Daily HRV & RHR was measured by the WHOOP® strap using reflectance photoplethysmography.^16^ RHR was measured in beats per minute (bpm) and HRV was measured in milliseconds (ms). HRV is calculated by the root-mean-square difference of successive heartbeat intervals.^7^ Improved cardiovascular fitness is indicated by higher HRV scores and lower RHR scores.

*Recorded Daily Activity* was measured by the WHOOP® strap by a three-axis accelerometer and processed using a proprietary algorithm to create daily activity records.

*Time Spent in Heart Rate Zones* was calculated by the time individuals spent in any of the six heart rate zones: Zone 0 = 0-50% heart rate reserve (HRR); zone 1 (50-60% HRR); zone 2 (60-70% HRR); zone 3 (70-80% HRR); zone 4 (80-90% HRR); and zone 5 (90-100% HRR).

These zones were measured automatically by the WHOOP® strap during exercise.^7^ This maximum heart rate zone was calculated at WHOOP® strap set up based on age, sex, and anthropometric measures entered by the participant.

*Daily strain* was measured using a proprietary formula and provided by the WHOOP® strap. Strain is a summary metric of the cardiovascular load, or the level of strain training takes on the cardiovascular system based on caloried burned, average heart rate, and max heart rate over the course of the day. Strain is scored on a scale from 0 to 21, with higher scores indicating more strenuous activity during the day that puts stress on the body.

*Recorded activities* were merged into a day-by-day measure of total time by day spent in each activity, and the three tables were merged by user ID and date. Daily minutes spent in a recorded activity in any zone were included as daily activity minutes, and daily minutes of zone 3 heart rate and higher were converted into a daily moderate/vigorous minutes variable. These tables were connected to the women’s pertinent pregnancy dates, including conception and delivery date. After calculating time to date variables, these dates were stripped from the data and not otherwise utilized.

## Data Analysis

Data analyses were conducted in SAS 9.4.^17^ Descriptive statistics are reported as frequencies and valid percentages of categorical variables, and mean, standard deviation, minimum and maximum values for continuous variables. Data were summarized by participant and by week for some descriptive analysis. Linear mixed models were used to model the longitudinal data. After assumptions were checked and found satisfactory, a variety of models were tested, including random intercept, random slope, both random intercept and slope, along with continuous (e.g., day to delivery) and categorical (e.g., trimester) time effects. Time was restricted to 43 weeks prior to birth and 8 weeks post-partum for the models, as most participants had data for this time-period. Splines were fitted for non-linear patterns with the continuous time fixed effects. The best fitting models were selected via lowest Akaike information criterion (AIC). Kuder-Richardson degree of freedom correction was used for all models. All available data was used via Restricted Maximum Likelihood (REML) method. The best fitting model included a random intercept and random continuous time slope (days to delivery), with a variance components covariance matrix and two splines set at different points for the two outcomes. A series of three models are presented for each of the two outcomes: Model 1: days until delivery only; Model 2: moderators for total activity minutes per day; Model 3: moderators for moderate/vigorous activity minutes per day. Fixed effects estimates along with standard errors, df, t-value and p-value are presented for each model.

## Results

### Participants

A total of 38 participants were enrolled. Eight participants withdrew from the study citing one of the following reasons: 1) no longer attempting conception, 2) not finding the wearable comfortable, or 3) feelings of additional stress of infertility. Of the 30 participants who continued to wear the strap during the study period, 12 did not conceive during the study period. Birth data was available for 18 participants. Women were followed between 142 and 754 days, with an average of 405.83 days (SD = 153.71), and total of 7,305 days logged. Based on monthly surveys, women had few medical conditions and were physically active throughout pregnancy.

All women gave birth between 37 and 41 weeks of pregnancy.

### Changes during Pregnancy

Based on WHOOP® data, activity levels decreased over the course of the pregnancy from almost 28 minutes of logged daily activity pre-pregnancy to 14 minutes by the third trimester (Table 1). There were also decreases in HRV and increases in RHR by trimester. Logged number of daily activity minutes was strongly correlated with moderate/vigorous daily activity minutes (r=0.82, p < 0.0001), and both indicators were correlated with the daily available strain score (activity r = 0.42, p < 0.0001; mod/vig r = 0.56, p < 0.0001).

**Table 1:**
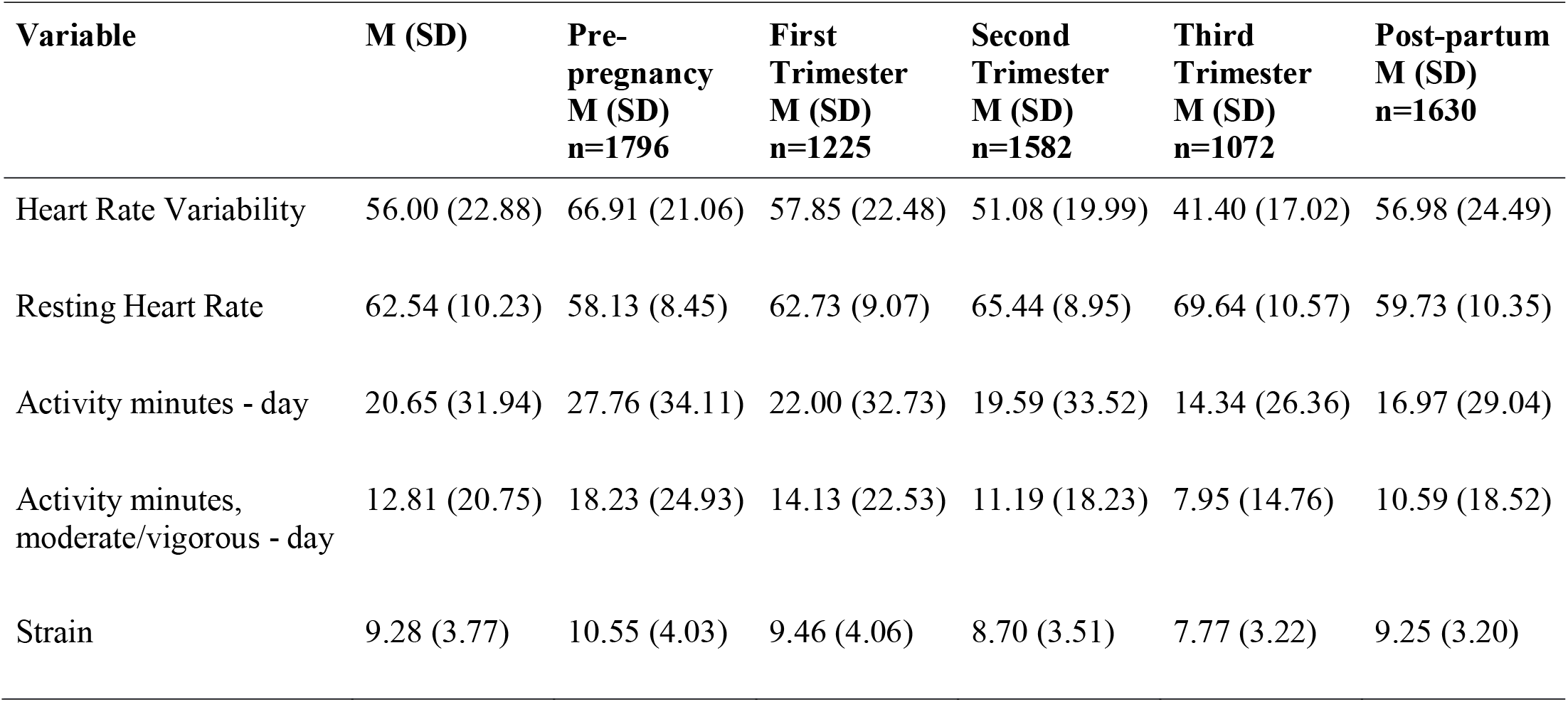
Descriptive statistics for variables of interest; n=18 across 7,305 days of data.

| **Variable** | **M (SD)** | **Pre-pregnancy**<br>**M (SD)**<br>**n=1796** | **First Trimester**<br>**M (SD)**<br>**n=1225** | **Second Trimester**<br>**M (SD)**<br>**n=1582** | **Third Trimester**<br>**M (SD)**<br>**n=1072** | **Post-partum**<br>**M (SD)**<br>**n=1630** |
| --- | --- | --- | --- | --- | --- | --- |
| Heart Rate Variability | 56.00 (22.88) | 66.91 (21.06) | 57.85 (22.48) | 51.08 (19.99) | 41.40 (17.02) | 56.98 (24.49) |
| Resting Heart Rate | 62.54 (10.23) | 58.13 (8.45) | 62.73 (9.07) | 65.44 (8.95) | 69.64 (10.57) | 59.73 (10.35) |
| Activity minutes - day | 20.65 (31.94) | 27.76 (34.11) | 22.00 (32.73) | 19.59 (33.52) | 14.34 (26.36) | 16.97 (29.04) |
| Activity minutes, moderate/vigorous - day | 12.81 (20.75) | 18.23 (24.93) | 14.13 (22.53) | 11.19 (18.23) | 7.95 (14.76) | 10.59 (18.52) |
| Strain | 9.28 (3.77) | 10.55 (4.03) | 9.46 (4.06) | 8.70 (3.51) | 7.77 (3.22) | 9.25 (3.20) |

Cardiovascular fitness decreased throughout pregnancy until 7-8 weeks prior to delivery, and then rapidly improved through and post-birth (Figures 1 and 2). A small reduction in HRV and increase in RHR can be seen in Figures 1 and 2 around the point of conception.

**Figure 1:**
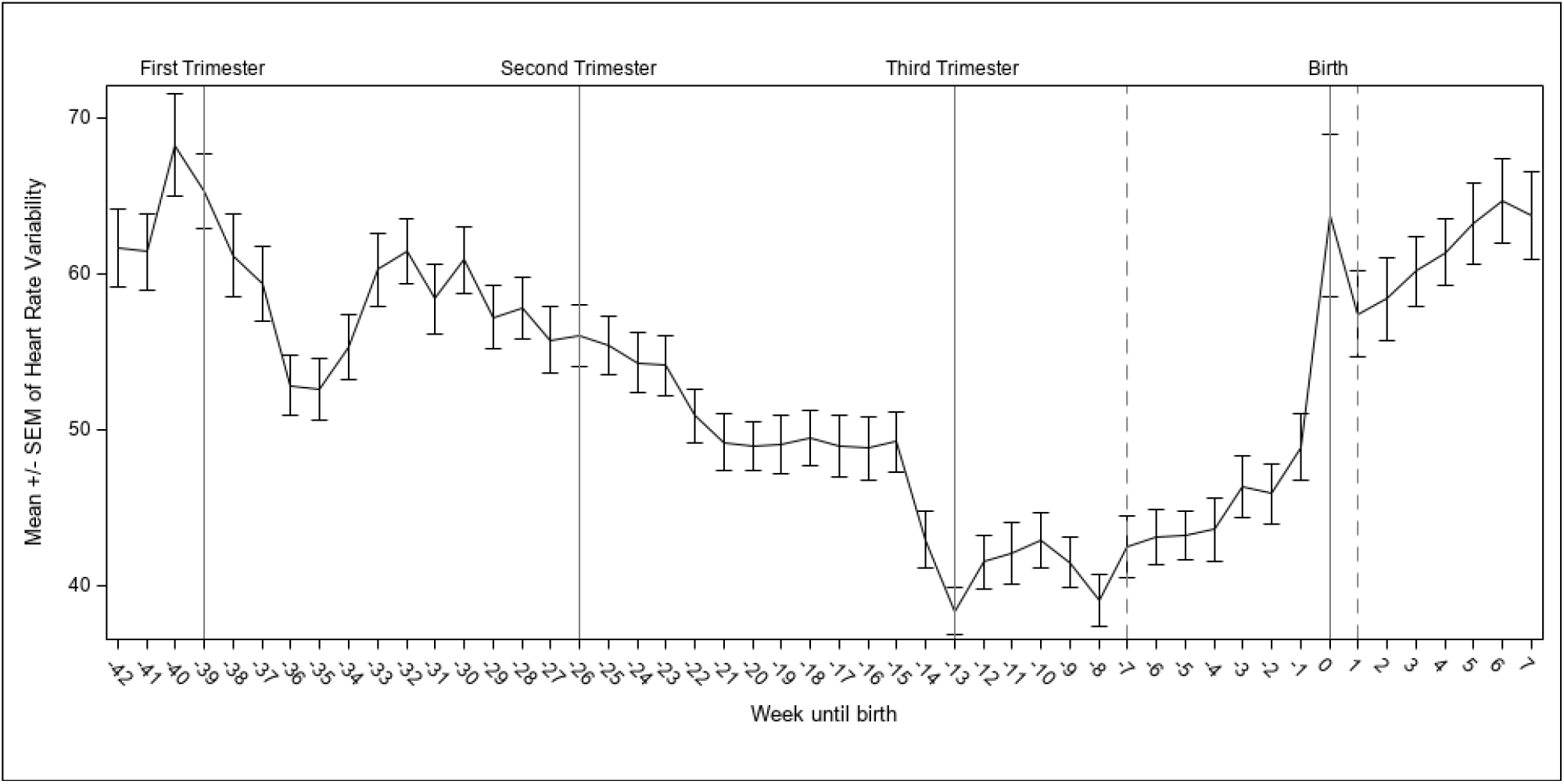
Weeks until birth, with splines indicated by a dotted line and referent lines included for the starts of trimester of pregnancy for average heart rate variability (n=18).

**Figure 2:**
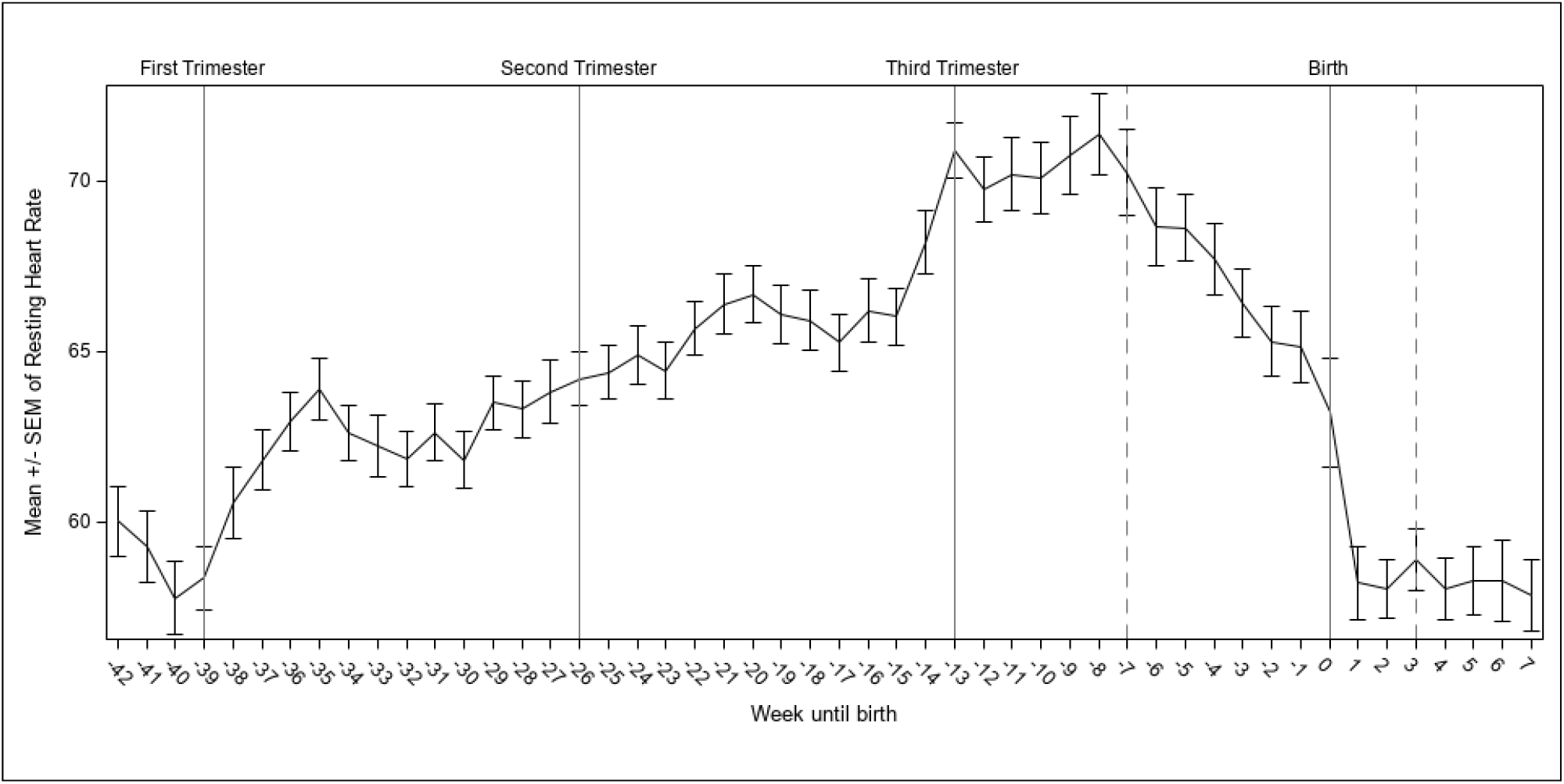
Weeks until birth, with splines indicated by a dotted line and referent lines included for the starts of trimester of pregnancy for average resting heart rate (n=18).

### Model Results for HRV and RHR

The linear mixed model results are presented for HRV (Table 2) and for RHR (Table 3). For both, the best fitting model is Model 2, which includes the moderating effects over time for daily logged activity minutes. Results generally attenuated for the moderate/vigorous daily activity minutes (i.e., Model 3 results). For HRV Model 2, there was a positive effect of activity on HRV (Est. = 0.07, p = 0.002). After accounting for activity, HRV decreased daily (Est. = - 0.10, p < 0.0001) until 49 days prior to delivery. Then there was a sharp increase in HRV daily (Est. = 0.38, p < 0.0001) until eight days post-partum, where HRV leveled off (Est. = -0.06, p = 0.30). Activity moderated the impact of early pregnancy on HRV (Est. = 0.0003, p = 0.02) until 49 days prior to delivery. It appeared to then slightly worsen the rapid improvement seen in HRV during the third trimester (Est. = -0.001, p = 0.003) but then improved HRV during post-partum (Est. = 0.005, p = 0.006).

**Table 2:**
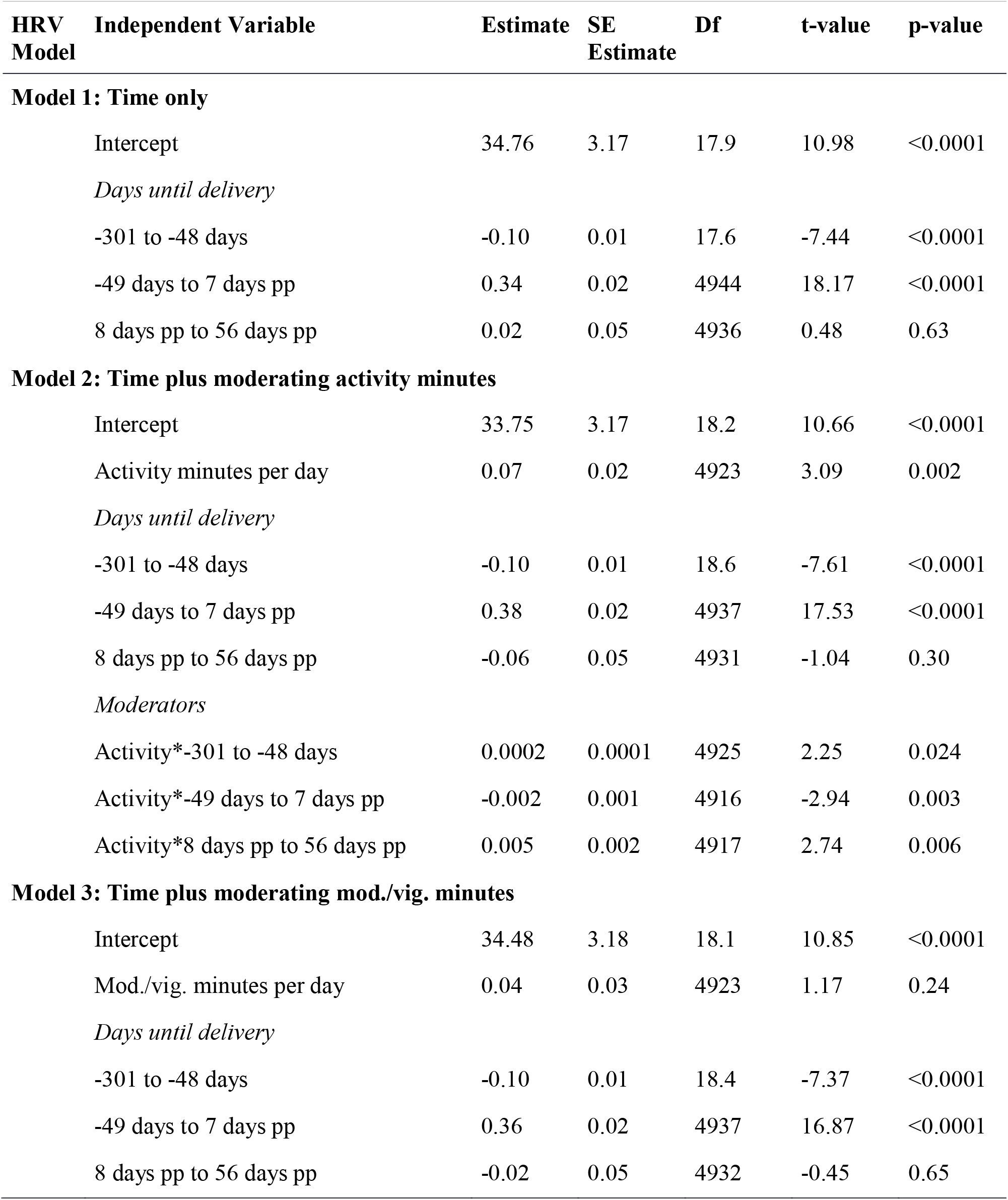
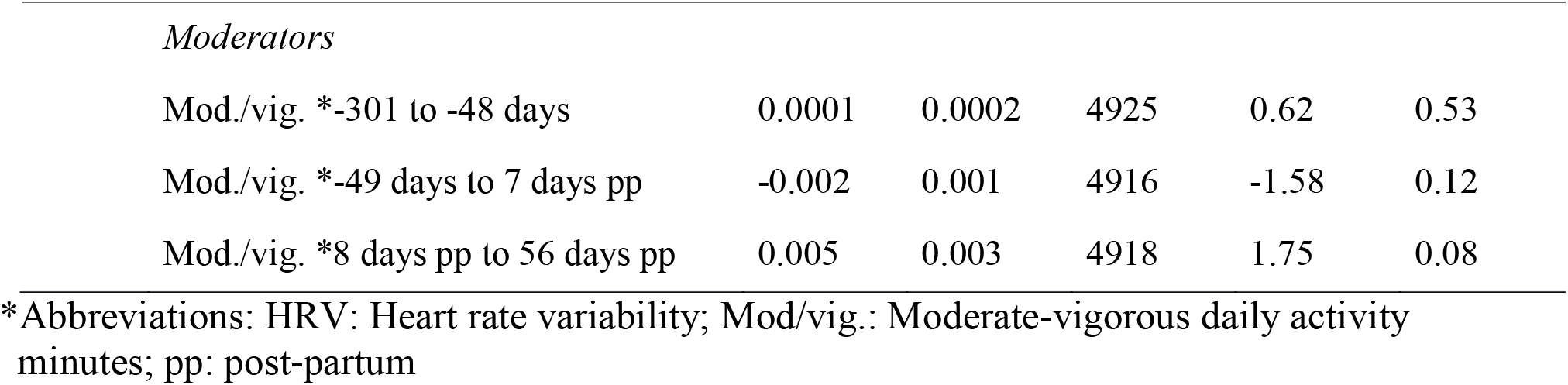
Fixed effects parameter estimates from linear mixed models for heart rate variability (HRV; n=18). Model 1: time only. Model 2: moderating effects of activity minutes per day. Model 3: moderating effects of moderate/vigorous activity minutes per day. Time was restricted to 43 weeks prior to birth and 8 weeks post-partum for the models.

| HRV Model | Independent Variable | Estimate | SE Estimate | Df | t-value | p-value |
| --- | --- | --- | --- | --- | --- | --- |
| **Model 1: Time only** |  |  |  |  |  |  |
|  | Intercept | 34.76 | 3.17 | 17.9 | 10.98 | <0.0001 |
|  | *Days until delivery* |  |  |  |  |  |
|  | -301 to -48 days | -0.10 | 0.01 | 17.6 | -7.44 | <0.0001 |
|  | -49 days to 7 days pp | 0.34 | 0.02 | 4944 | 18.17 | <0.0001 |
|  | 8 days pp to 56 days pp | 0.02 | 0.05 | 4936 | 0.48 | 0.63 |
| **Model 2: Time plus moderating activity minutes** |  |  |  |  |  |  |
|  | Intercept | 33.75 | 3.17 | 18.2 | 10.66 | <0.0001 |
|  | Activity minutes per day | 0.07 | 0.02 | 4923 | 3.09 | 0.002 |
|  | *Days until delivery* |  |  |  |  |  |
|  | -301 to -48 days | -0.10 | 0.01 | 18.6 | -7.61 | <0.0001 |
|  | -49 days to 7 days pp | 0.38 | 0.02 | 4937 | 17.53 | <0.0001 |
|  | 8 days pp to 56 days pp | -0.06 | 0.05 | 4931 | -1.04 | 0.30 |
|  | *Moderators* |  |  |  |  |  |
|  | Activity*-301 to -48 days | 0.0002 | 0.0001 | 4925 | 2.25 | 0.024 |
|  | Activity*-49 days to 7 days pp | -0.002 | 0.001 | 4916 | -2.94 | 0.003 |
|  | Activity*8 days pp to 56 days pp | 0.005 | 0.002 | 4917 | 2.74 | 0.006 |
| **Model 3: Time plus moderating mod./vig. minutes** |  |  |  |  |  |  |
|  | Intercept | 34.48 | 3.18 | 18.1 | 10.85 | <0.0001 |
|  | Mod./vig. minutes per day | 0.04 | 0.03 | 4923 | 1.17 | 0.24 |
|  | *Days until delivery* |  |  |  |  |  |
|  | -301 to -48 days | -0.10 | 0.01 | 18.4 | -7.37 | <0.0001 |
|  | -49 days to 7 days pp | 0.36 | 0.02 | 4937 | 16.87 | <0.0001 |
|  | 8 days pp to 56 days pp | -0.02 | 0.05 | 4932 | -0.45 | 0.65 |

|  |  |  |  |  |  |
| --- | --- | --- | --- | --- | --- |
| *Moderators* |  |  |  |  |  |
| Mod./vig. *-301 to -48 days | 0.0001 | 0.0002 | 4925 | 0.62 | 0.53 |
| Mod./vig. *-49 days to 7 days pp | -0.002 | 0.001 | 4916 | -1.58 | 0.12 |
| Mod./vig. *8 days pp to 56 days pp | 0.005 | 0.003 | 4918 | 1.75 | 0.08 |

\*Abbreviations: HRV: Heart rate variability; Mod/vig.: Moderate-vigorous daily activity minutes; pp: post-partum

**Table 3:**
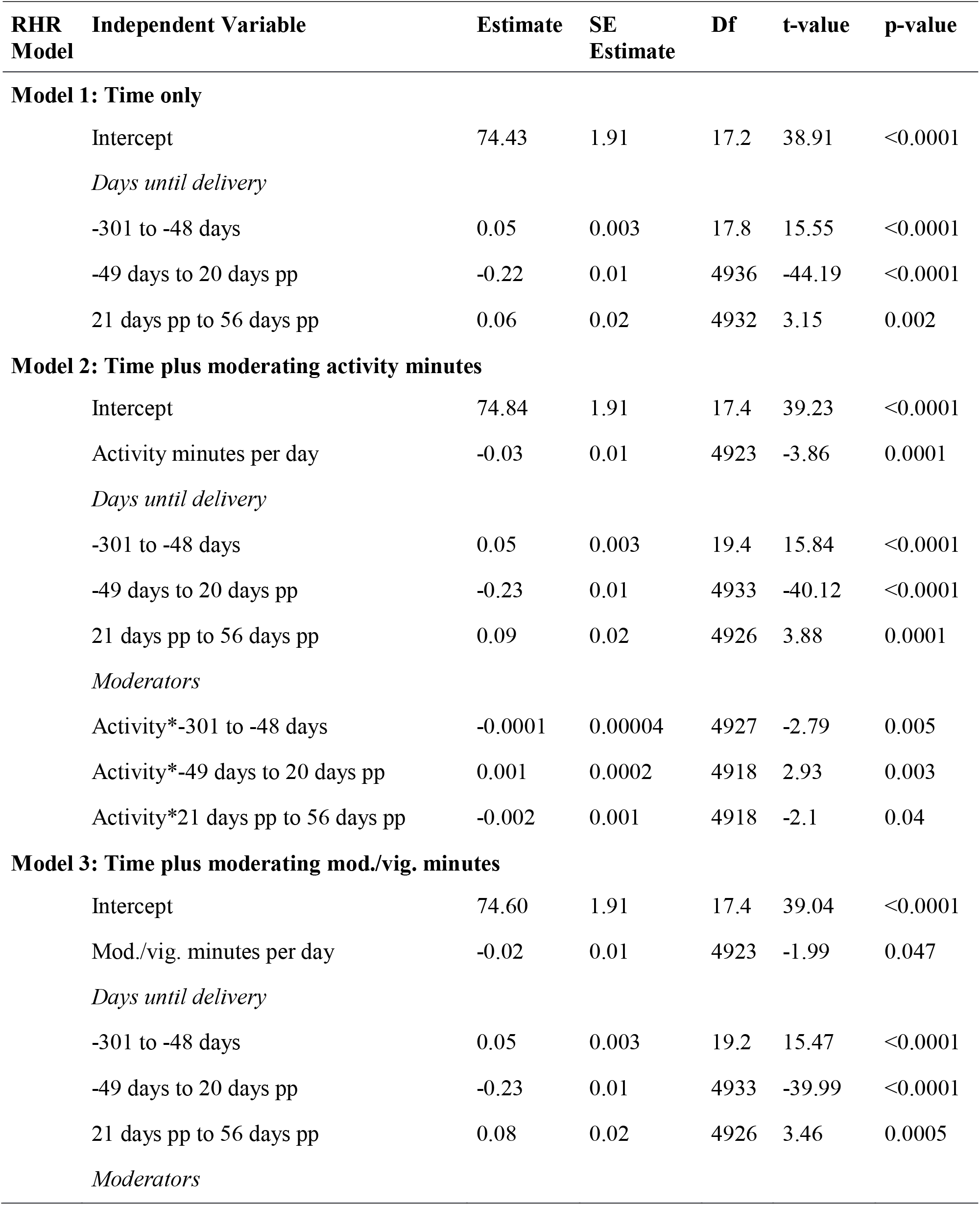
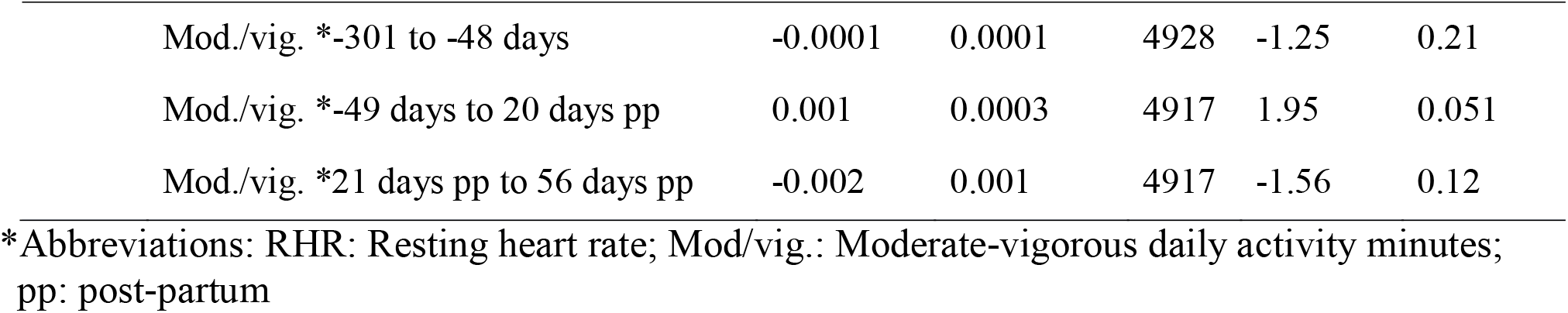
Fixed effects parameter estimates from linear mixed models for resting heart rate (RHR, n=18). Model 1: time only. Model 2: moderating effects of activity minutes per day. Model 3: moderating effects of moderate/vigorous activity minutes per day. Time was restricted to 43 weeks prior to birth and 8 weeks post-partum for the models.

| **RHR Model** | **Independent Variable** | **Estimate** | **SE Estimate** | **Df** | **t-value** | **p-value** |
| --- | --- | --- | --- | --- | --- | --- |
| **Model 1: Time only** |  |  |  |  |  |  |
|  | Intercept | 74.43 | 1.91 | 17.2 | 38.91 | <0.0001 |
|  | *Days until delivery* |  |  |  |  |  |
|  | -301 to -48 days | 0.05 | 0.003 | 17.8 | 15.55 | <0.0001 |
|  | -49 days to 20 days pp | -0.22 | 0.01 | 4936 | -44.19 | <0.0001 |
|  | 21 days pp to 56 days pp | 0.06 | 0.02 | 4932 | 3.15 | 0.002 |
| **Model 2: Time plus moderating activity minutes** |  |  |  |  |  |  |
|  | Intercept | 74.84 | 1.91 | 17.4 | 39.23 | <0.0001 |
|  | Activity minutes per day | -0.03 | 0.01 | 4923 | -3.86 | 0.0001 |
|  | *Days until delivery* |  |  |  |  |  |
|  | -301 to -48 days | 0.05 | 0.003 | 19.4 | 15.84 | <0.0001 |
|  | -49 days to 20 days pp | -0.23 | 0.01 | 4933 | -40.12 | <0.0001 |
|  | 21 days pp to 56 days pp | 0.09 | 0.02 | 4926 | 3.88 | 0.0001 |
|  | *Moderators* |  |  |  |  |  |
|  | Activity*-301 to -48 days | -0.0001 | 0.00004 | 4927 | -2.79 | 0.005 |
|  | Activity*-49 days to 20 days pp | 0.001 | 0.0002 | 4918 | 2.93 | 0.003 |
|  | Activity*21 days pp to 56 days pp | -0.002 | 0.001 | 4918 | -2.1 | 0.04 |
| **Model 3: Time plus moderating mod./vig. minutes** |  |  |  |  |  |  |
|  | Intercept | 74.60 | 1.91 | 17.4 | 39.04 | <0.0001 |
|  | Mod./vig. minutes per day | -0.02 | 0.01 | 4923 | -1.99 | 0.047 |
|  | *Days until delivery* |  |  |  |  |  |
|  | -301 to -48 days | 0.05 | 0.003 | 19.2 | 15.47 | <0.0001 |
|  | -49 days to 20 days pp | -0.23 | 0.01 | 4933 | -39.99 | <0.0001 |
|  | 21 days pp to 56 days pp | 0.08 | 0.02 | 4926 | 3.46 | 0.0005 |
|  | *Moderators* |  |  |  |  |  |

|  |  |  |  |  |  |
| --- | --- | --- | --- | --- | --- |
| Mod./vig. *-301 to -48 days | -0.0001 | 0.0001 | 4928 | -1.25 | 0.21 |
| Mod./vig. *-49 days to 20 days pp | 0.001 | 0.0003 | 4917 | 1.95 | 0.051 |
| Mod./vig. *21 days pp to 56 days pp | -0.002 | 0.001 | 4917 | -1.56 | 0.12 |

\*Abbreviations: RHR: Resting heart rate; Mod/vig.: Moderate-vigorous daily activity minutes; pp: post-partum

RHR followed a similar pattern, with a favorable effect of activity (Est. = -0.03, p = 0.0001). After accounting for activity, RHR increased daily (Est. = 0.05, p < 0.0001) until 49 days prior to delivery. Then there was a sharp decrease in RHR daily (Est. = -0.23, p < 0.0001) until 21 days post-partum, where RHR returned to pre-pregnancy levels (Est. = 0.08, p = 0.0001). Activity moderated the impact of early pregnancy on RHR (Est. = -0.0001, p = 0.005) until 49 days prior to delivery. It appeared to then slightly worsen the rapid improvement seen in RHR during the third trimester (Est. = 0.001, p = 0.003) but then improved RHR during post-partum (Est. = -0.002, p = 0.04).

## Discussion

The 2018 U.S. Department of Health and Human Services Physical Activity Guidelines for Americans state that women who engage in high-intensity aerobic activity or are otherwise physically active prior to pregnancy can continue these activities during pregnancy and into the postpartum period. The report also recommends that women exercise at least 150 minutes per week during pregnancy and in postpartum.^18^ Physical activity during pregnancy is associated with lower HR and higher HRV in both the mothers and the fetus, when compared to pregnant women who are not physically active,^19, 20^ along with increased stroke volume and increased oxygen uptake.^21, 22^ Previous research has shown that RHR increases by 3-5% during the first semester, 10-15% during the second trimester, and 15-20% in the third trimester, and returns to pre-pregnancy values within 3-6 months postpartum. ^1^

In this study, we used continuous monitoring during pregnancy as well as pre-pregnancy and postpartum data to define the physiologic changes that occur with HRV and RHR. As defined in previous studies, HRV decreased per trimester when reviewed as a whole.^13, 23^

However, in this sample of healthy and active women, cardiovascular health generally decreased over the course of the pregnancy until roughly 49 days (7 weeks) prior to birth. At that time, cardiovascular health indicators rapidly improved until post birth even beyond pre-pregnancy levels.

In this sample of generally active women, more activity minutes per day mitigated some of the negative impact of pregnancy on cardiovascular health and helped with improvements post-pregnancy. The slight worsening in cardiovascular health in the 49 days prior to birth through the date of birth may be related to the participants’ improved cardiovascular health prior to that point. This effect was stronger for general activity minutes than for moderate/vigorous minutes, suggesting the amount of activity may be more beneficial than the type of activity.

Currently, obstetricians do not consistently provide clear exercise recommendations for their patients, especially those who are sedentary (McGee et al., 2018). The findings of this study indicate any type of activity is beneficial to overall cardiovascular health during pregnancy.

There are significant clinical implications for these findings due to the ease of integrating additional minutes of activity rather than increasingly rigorous or different types of exercise. Obstetrician’s most limited exercise recommendations have been on resistance training, maximum heart rate during exercise, and third trimester exercise (McGee et al., 2018), so this allows for easier recommendations to be disseminated while still improving patient’s cardiovascular health.

### Strengths and Limitations

This study has several considerable strengths. Although only 18 participants were analyzed, due to the large amount of data that was able to be obtained from the WHOOP® straps, this allowed for substantial statistical power despite the cohort size. Second, the use of the WHOOP® strap resulted in daily readings, in comparison to other studies which obtained readings at discrete points during pregnancy. Third, to the best of our knowledge, this is the first study to use continuous monitoring during pregnancy of HRV and RHR. Additionally, we were able to follow the participants prior to conception, for the duration of their pregnancies and into postpartum, for an average duration of 405.8±153 days. This allowed us to get in-depth and continuous insight into these different times of the perinatal experience for a considerable length of time. Finally, all the participants in this study delivered at term with no reported pregnancy complications suggesting this data set can be representative of normal healthy pregnancies.

Limitations include the small cohort of patients, many of whom were seeking infertility treatment. Although there may be a possible lack of generalizability outside the cohort, the sample size was sufficient for detecting significant changes over the perinatal period for HRV and RHR.

## Conclusions

In summary, this data has strong implications for all pregnant women. In contrast to previous literature that examined cardiovascular health at discrete timepoints, our daily data demonstrates that there is a sharp improvement in cardiovascular health in all women with uncomplicated term pregnancies prior to birth. Consistent with other data, but in extensive detail, we demonstrate that pregnant women who were able to be more active had consistently improved RHR and HRV particularly during the first few trimesters of pregnancy and during the post-pregnancy recovery period. Using this data from uncomplicated term pregnancies, future studies can evaluate HRV and RHR in women who are not currently active as well as in women at high risk for complications.

## Data Availability

All data produced in the present study are available upon reasonable request to the authors

